# Interpretation of non-responders to SARS-CoV-2 vaccines using WHO International Standard

**DOI:** 10.1101/2022.03.31.22273272

**Authors:** Qiang Zeng, Xue Yang, Qi Gao, Biao-yang Lin, Yong-zhe Li, Gang Huang, Yang Xu

## Abstract

Severe acute respiratory syndrome coronavirus 2 (SARS-CoV-2) has caused a global pandemic with more than 485 millions infected. Questions about non-responders to SARS-CoV-2 vaccines remain unaddressed. Here, we report data from people after administering the complete dose of SARS-CoV-2 vaccines using the World Health Organization International Standard for anti-SARS-CoV-2 immunoglobulin. Our study showed that immune cells such as CD4 cells, CD8 cells, and B cells and anti-spike immunoglobulin G levels were significantly reduced in the elderly. There were 7.5% non-responders among the 18–59 yr group and 11.7% in the ≥60 yr group. A titer of anti-SARS-CoV-2 spike immunoglobulin G is blew 50 BAU/mL to be considered as non-responders at intervals of 30 to 90 days after the last vaccine dose. Booster vaccination may be recommended for non-responders to reduce the disease severity and mortality.

## INTRODCTION

Severe acute respiratory syndrome coronavirus 2 (SARS-CoV-2) has caused a global pandemic, infecting more than 485 million people and killing more than 6 million.^1^ Since December 2020 the World Health Organization (WHO) recommends vaccination against COVID-19, nine types of coronavirus disease 2019 (COVID-19) vaccines have been included in the emergency use list.^2^

Vaccination against COVID-19 is especially important in reducing severe illness and mortality. According to the data of the Centers for Disease Control and Prevention (CDC) in 2016-2017, the mortality rate caused by influenza virus was 0.13%.^3^

In order to bring the COVID-19 pandemic under control as soon as possible and ensure that the mortality rate of COVID-19 is close to that caused by influenza virus, the prevention and treatment of children as well as elderly and immunocompromised people has emerged as a top priority at present.^4-8^ Sun et al. first reported that hospitalization and severe outcomes were similar in unvaccinated healthy individuals and immunocompromised patients who received full SARS-CoV-2 vaccination in the United States, suggesting that COVID-19 breakthrough infection after SARS-CoV-2 vaccination is associated with immune dysfunction. Hospitalization and severe outcomes were 21.1% and 1.9%, respectively, in unvaccinated healthy individuals, and 20.7% and 2.1%, respectively, in patients with immune dysfunction after 14 days following full vaccination, indicating that an immune barrier is not well established in immunocompromised patients after full vaccination and post-vaccination testing is necessary to identify immunocompromised individuals without specific immunity so they can be given additional prophylaxis after full vaccination.^8^ This study suggests that post-vaccination testing will help reduce mortality, showing the importance and urgency of post-vaccination assessments using an international standard.

To date, more than 5 billion people have been vaccinated against COVID-19.^9^ In clinical trials associated with COVID-19 vaccines, the effective COVID-19 vaccination reportedly elicits specific antibody responses. An effective humoral immune response is defined as a ≥ 4-fold increase in antibody titers from baseline within 1–3 months of the vaccination procedure and is considered gold standard for assessing antibody protection in vaccinated recipients.^10-12^ In contrast, a non-responder is an individual who demonstrates no effective humoral immune response despite the completion of the suggested vaccination procedure.^13-14^

During the promotion of vaccination, several factors affecting the response to the COVID-19 vaccines were taken into consideration, especially the reduced response to the COVID-19 vaccine in children, elderly people, and immunocompromised population. However, despite the completion of COVID-19 vaccination in the population as per the recommendations by WHO, the outcomes concerning the protective levels of antibody concentration and factors determining the identification of non-responders still remain inconclusive.^15-25^

To this end, in December 2020, WHO issued an international standard (IS) for the quantification of anti-SARS-CoV-2 immunoglobulin for post-vaccination testing.^26-27^ This standard provides a unified benchmark for effective antibody protective concentrations after vaccination. In this clinical study, we analyzed 627 people that volunteered to participate in COVID-19 vaccination and subsequent assessment of antibody titers. After two doses of vaccination, the antibody titer was evidently increased by ≥ 4 times from baseline as the gold standard. Furthermore, the data using the WHO IS was comprehensively analyzed to provide insights for improving the efficacy of vaccines, help in reduction of breakthrough infections after vaccines, and ultimately to reduce the disease severity and mortality.

## RESULTS

### Immune characteristics of 627 cases prior to vaccination

There were 42.4% (266/627) individuals aged ≥60 yr and 50.9% (319/627) male enrolled in the study (Table 1). In the 18–59 yr group, the medians [interquartile ranges (IQRs)] of ALC, CD4 cell count, CD8 cell count, B cell count, and NK cell count were 1,476 (1,168–1,875), 851 (677– 1,151), 490 (357–632), 256 (179–367), and 193 (141–287)/mm^3^, respectively. On the contrary, in the ≥60 yr group, the respective medians (IQRs) were 1,281 (1,023–1,520), 747 (562–955), 418 (288–544), 204 (138–303), and 234 (162–355)/mm^3^ (Table 1). In fact, the number of naïve lymphocytes, CD4 cells, CD8 cells, and B cells were significantly reduced in the elderly population than that in the 18–59 yr population (*P* < 0.001). Hence, these naïve immune cells wane significantly, while the NK cell counts increase significantly in the elderly people (Table 1, Fig. 1A).

**Table 1.**
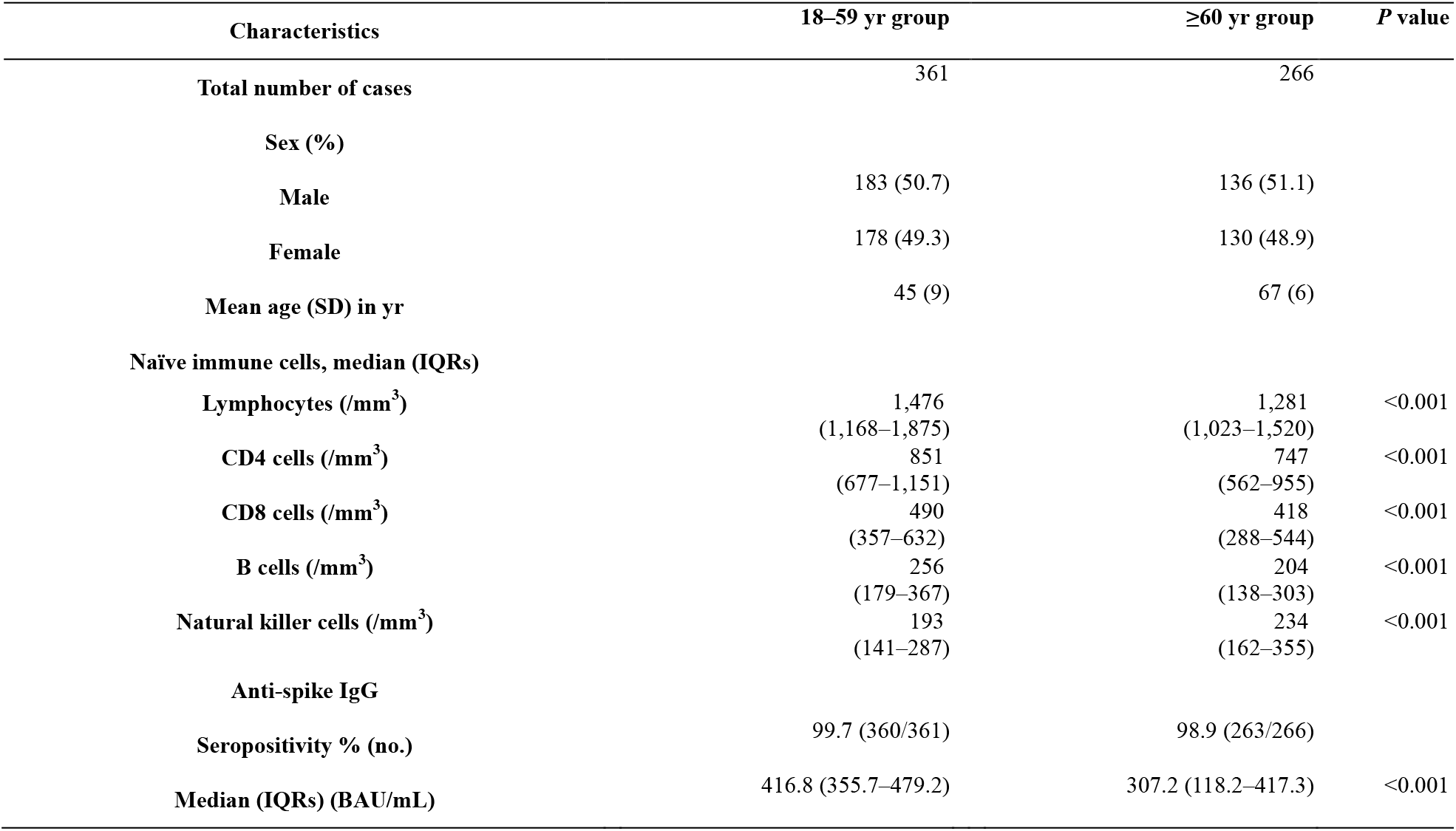
Immune characteristics of individuals before and after vaccination.

| Characteristics | 18–59 yr group | ≥60 yr group | *P* value |
| --- | --- | --- | --- |
| **Total number of cases** | 361 | 266 |  |
| **Sex (%)** |  |  |  |
| **Male** | 183 (50.7) | 136 (51.1) |  |
| **Female** | 178 (49.3) | 130 (48.9) |  |
| **Mean age (SD) in yr** | 45 (9) | 67 (6) |  |
| **Naïve immune cells, median (IQRs)** |  |  |  |
| **Lymphocytes (/mm<sup>3</sup>)** | 1,476<br>(1,168–1,875) | 1,281<br>(1,023–1,520) | <0.001 |
| **CD4 cells (/mm<sup>3</sup>)** | 851<br>(677–1,151) | 747<br>(562–955) | <0.001 |
| **CD8 cells (/mm<sup>3</sup>)** | 490<br>(357–632) | 418<br>(288–544) | <0.001 |
| **B cells (/mm<sup>3</sup>)** | 256<br>(179–367) | 204<br>(138–303) | <0.001 |
| **Natural killer cells (/mm<sup>3</sup>)** | 193<br>(141–287) | 234<br>(162–355) | <0.001 |
| **Anti-spike IgG** |  |  |  |
| **Seropositivity % (no.)** | 99.7 (360/361) | 98.9 (263/266) |  |
| **Median (IQRs) (BAU/mL)** | 416.8 (355.7–479.2) | 307.2 (118.2–417.3) | <0.001 |

**Fig. 1.**
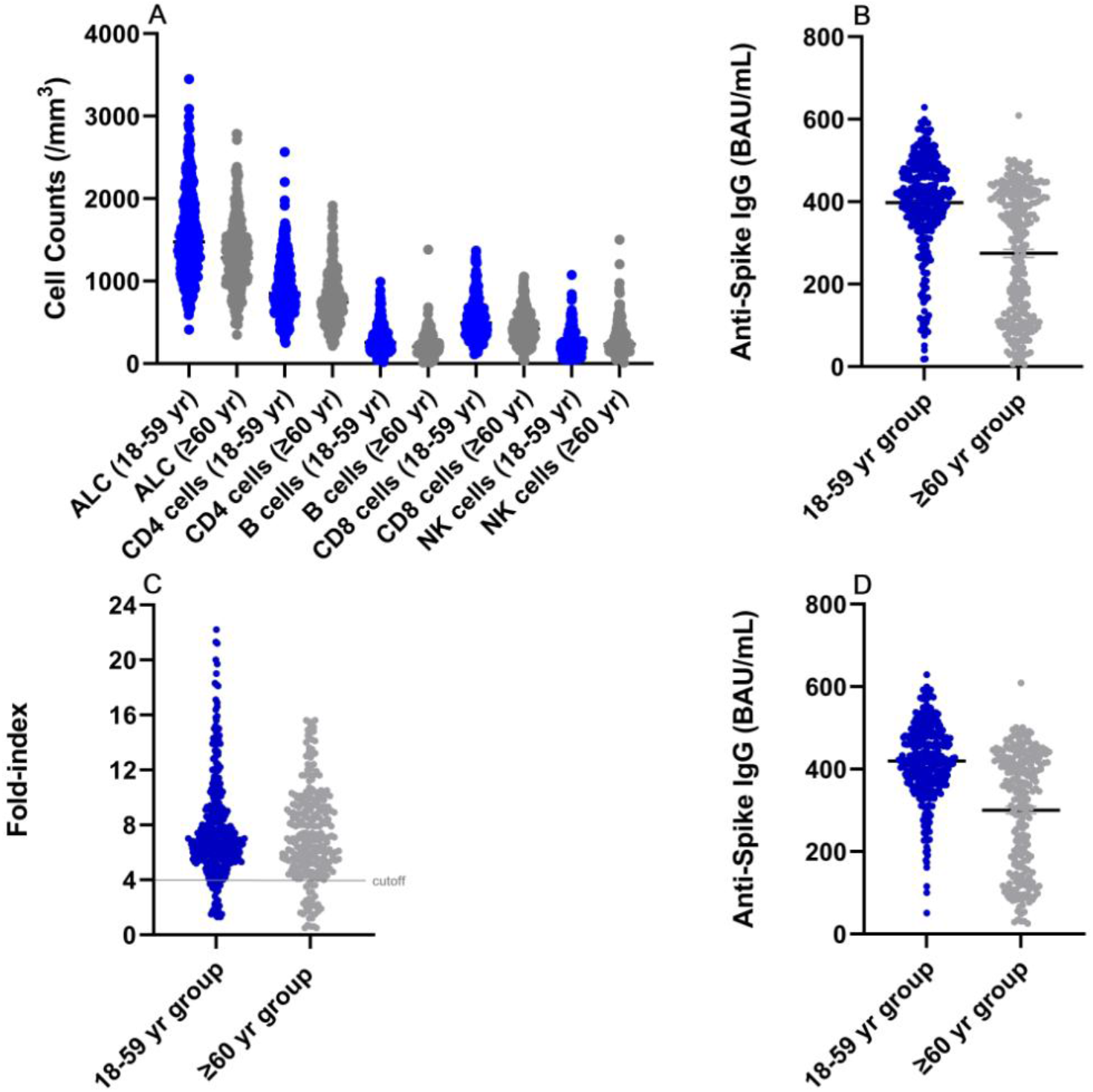
Immune characteristics of 627 individuals. (A) Naïve cellular immune parameters of the 627 cases who received physical examinations. These naïve immune cells wane significantly, while the natural killer (NK) cell counts increase significantly in the elderly people. ALC = absolute lymphocyte count. (B) The anti-spike IgG levels after complete vaccination of the 627 cases. The quantitative level of the anti-spike IgG was significantly lower in the ≥60 yr group (median 307.2, IQRs 118.2–417.3 BAU/mL) than that in the 18–59 yr group (median 416.8, IQRs 355.7–479.2 BAU/mL, *P* < 0.001. Mean and standard error of the mean (SEM) were shown. (C) The vaccine-induced responses using the 4-fold increase after complete vaccination of the 627 cases. There were 7.5% non-responders (fold-index < 4) among the 18–59 yr group and 11.7% in the ≥60 yr group. The reference ranges (1–99 percentile) for responders (fold–index ≥ 4) were 43.9–592.0 BAU/mL in combination of the 18–59 yr group and the ≥60 yr group. A cutoff line at fold-index 4 was showed. (D) In the responder group (fold-index ≥ 4), intervals for 1– 99 percentile were 131.8–592.3 BAU/mL in the 18–59 yr group, and 29.7–500.9 BAU/mL in the ≥60 yr group, respectively. Mean and standard error of the mean (SEM) were shown.

### Anti-spike IgG levels in the 627 cases after complete vaccination

We analyzed the anti-spike immunoglobulin (Ig)G levels after complete vaccination of the 627 cases (Table 1). Post-vaccination testing was done at intervals of 14 to 90 days after the second vaccine dose. The anti-spike IgG seropositive rates were 99.7% in the 18–59 yr population and 98.9% in the ≥60 yr population based on the cutoff (Table 1). However, the quantitative level of the anti-spike IgG was significantly lower in the ≥60 yr group (median 307.2, IQRs 118.2–417.3 BAU/mL) than that in the 18–59 yr group (median 416.8, IQRs 355.7–479.2 BAU/mL) (Table 1, Fig. 1B). The reference ranges (2.5–97.5 percentile) were 88.6–576.2 BAU/mL in the 18–59 yr group and 27.7–491.0 BAU/mL in the ≥60 yr group at 14–90 days after complete vaccination (Table 2).

**Table 2.** Characteristics of seroconversion after the complete dose of inactivated SARS-CoV-2 vaccines.

| Characteristics | 18–59 yr group |  |  | ≥60 yr group |  |  |
| --- | --- | --- | --- | --- | --- | --- |
|  | Fold-index <4 | Fold-index ≥4 | *P* value | Fold-index <4 | Fold-index ≥4 | *P* value |
| **Total number of cases** | 361 |  |  | 266 |  |  |
| **Anti-spike IgG BAU/mL (2.5–97.5 percentile)** | 88.6–576.2 |  |  | 27.7–491.0 |  |  |
| **Fold-index % (no.)\*** | 7.5 (27/361) | 92.5 (334/361) |  | 11.7 (31/266) | 88.3 (235/266) |  |
| **Anti-spike IgG BAU/mL** |  |  |  |  |  |  |
| **Median (IQRs)** | 115.8 (88.6–167.8) | 420.8 (369.9–480.6) | <0.0001 | 63.9 (35.1–106.9) | 346.0 (160.4–424.7) | <0.0001 |
| **2.5–97.5 percentile** | 18.3–266.3 | 200.7–576.5 |  | 5.4–317.8 | 46.6–491.1 |  |
| **Naïve immune cells (/mm<sup>3</sup>)** |  |  |  |  |  |  |
| **Lymphocytes, mean (95% CI)** | 1,130 (1,007–1,252) | 1,578 (1,524–1,633) | <0.0001 | 1,015 (888–1,143) | 1,344 (1,291–1,397) | <0.0001 |
| **CD4 cells, mean (95% CI)** | 631 (555–708) | 942 (905–979) | <0.0001 | 563 (494–631) | 818 (777–858) | <0.0001 |
| **CD8 cells, mean (95% CI)** | 414 (349–479) | 532 (508–557) | 0.0081 | 394 (310–478) | 444 (420–468) | 0.1744 |
| **B cells, mean (95% CI)** | 119 (72–166) | 306 (289–323) | <0.0001 | 74 (60–88) | 248 (231–266) | <0.0001 |
| **NK cells, mean (95% CI)** | 192 (151–233) | 235 (220–251) | 0.1241 | 281 (225–337) | 286 (261–311) | 0.8902 |

\*Post-vaccination testing was done at intervals of 14 to 90 days after the second vaccine dose. The reference ranges were defined as the 2.5–97.5 percentile in the study.

### Characteristics of seroconversion after the complete dose

Thereafter, we evaluated the vaccine-induced responses, based on the post-second-dose and pre-second-dose titers, using the 4-fold increase parameter (fold-index <4 or ≥4) (Table 2). Remarkably, there were 7.5% non-responders (fold-index < 4) among the 18–59 yr group and 11.7% in the ≥60 yr group (Table 2, Fig. 1C), indicating that the positive rate of anti-spike IgG cannot represent the seroconversion rate. Therefore, the anti-spike IgG positivity or seroprevalence might not be a suitable predictor of seroconversion (fold–index ≥ 4).

In the 18–59 yr group, the median (IQRs) levels of anti-spike IgG and the reference ranges were 115.8 (88.6–167.8) and 18.3–266.3 BAU/mL with fold-index < 4, respectively and 420.8 (369.9– 480.6) and 200.7–576.5 BAU/mL with fold-index ≥ 4, respectively (*P* < 0.0001). In contrast, in the ≥60 yr group, the median (IQRs) levels of anti-spike IgG and the reference ranges were 63.9 (35.1–106.9) and 5.4–317.8 BAU/mL with fold-index < 4, respectively and 346.0 (160.4–424.7) and 46.6–491.1 BAU/mL with fold-index ≥ 4, respectively (*P* < 0.0001). The reference ranges (1–99 percentile) for responders (fold–index ≥ 4) were 43.9–592.0 BAU/mL in combination of the 18–59 yr group and the ≥60 yr group at 14–90 days after complete vaccination (Figure 1C).

We further observed that the seroconversion rate was significantly related to the proportion of certain naïve immune cells (Table 2). Particularly, the lymphocyte count was significantly different (*P* < 0.0001) between the fold-index <4 or ≥4 groups. For instance, in the 18–59 yr group, the lymphocyte count was 1,130/mm^3^ [95% CI (1,007–1,252)/mm^3^] in the <4 group and 1,578/mm^3^ [95% CI (1,524–1,633)/mm^3^] in the ≥4 group. On the contrary, in the ≥60 yr group, the lymphocyte count was 1,015/mm^3^ [95% CI (888–1,143)/mm^3^] in the <4 group and 1,344/mm^3^ [95% CI 1,291–1,397)/mm^3^] in the ≥4 group. Similarly, the CD4 cell counts were significantly different (*P* < 0.0001) between the individuals with fold–index <4 and ≥4. For instance, in the 18–59 yr population, the CD4 cell count was 631/mm^3^ [95% CI (555–708)/mm^3^] versus 942/mm^3^ [95% CI (905–979)/mm^3^] in the <4 and ≥4 groups, respectively, while in the ≥60 yr age group, it was 563/mm^3^ [95% CI (494–631)/mm^3^] versus 818/mm^3^ [95% CI (777– 858)/mm^3^] in the <4 and ≥4 groups, respectively. With respect to the B cell count, there was a significant difference (*P* < 0.0001) between the individuals with fold–index < 4 and ≥ 4. For example, in the 18–59 age group, the B cell count was 119/mm^3^ [95% CI (72–166)/mm^3^ versus 306/mm^3^ [95% CI (289–323)/mm^3^] in the <4 and ≥4 groups, respectively, whereas in the ≥60 yr age group, it was 74/mm^3^ [95% CI (60–88) /mm^3^] versus 248/mm^3^ [95% CI (231–266)/mm^3^] in the <4 and ≥4 groups, respectively. Regarding the CD8 cell count, a significant difference was noted only between the individuals with <4 and ≥4 fold-indices in the 18–59 yr age group [414/mm^3^ (95% CI 349–479/mm^3^) versus 532/mm^3^ (95% CI 508–557/mm^3^), *P* = 0.0081]. However, the CD8 cell count in the ≥60 yr group and NK cell count in both the age groups did not portray any significant differences.

## DISCUSSION

To the best of our knowledge, this is the first clinical study to report non-responders after administering the complete dose of inactivated SARS-CoV-2 vaccines using WHO International Standard (IS) for anti-SARS-CoV-2 immunoglobulin (Ig). Whether there is a humoral immune response following COVID-19 vaccination is a marker of population immunity.^28-30^ Typically, effective humoral immune response is defined as a ≥ 4-fold rise in antibody titers from baseline within 1-3 months of the vaccination schedule. The use of anti-SARS-CoV-2 assays with the WHO IS can facilitate the comparison of the strength of the humoral immune response between individuals, making the data more accurate and providing reliable data for the COVID-19 vaccine booster. Therefore, adequate clinical trials are necessary regarding the assessment of immune characteristics of individuals prior to vaccine booster shot, such that the mortality in the pandemic may be quickly reduced.

We used an anti-SARS-CoV-2 spike quantitative IgG kit (COVID-SeroKlir Kantaro SARS-CoV-2 IgG Ab Kit) approved by the Food and Drug Administration (FDA) under Emergency Use Authorization (EUA) with the WHO IS. This kit has been extensively evaluated in many clinical studies, including neutralizing antibodies after SARS-CoV-2 infection, immunological memory to SARS-CoV-2, convalescent plasma treatment of severe COVID-19, and antibody responses to mRNA vaccines in healthy people and patients.^30-35^ After complete two dose vaccination, the reference ranges (1–99 percentile) for all responders (fold–index ≥ 4) were 43.9–592.0 BAU/mL. A preliminary cutoff of 50 BAU/mL was set based on percentiles of all responders and convenience of manufacturing standard controls. The final cutoff value will be determined by future clinical trials.

The WHO IS has demonstrated to be enabled to comparison between different types of vaccines. Zitt et al. reported that the median titers of non-seroconversion and seroconversion were 635.5 and 1,565.0 BAU/mL after two doses of mRNA vaccination in hemodialysis patients at 67.6 ± 14.8 years, respectively;^36^ whereas we reported that the median titers of non-seroconversion and seroconversion were 63.9 and 346.0 BAU/mL after giving two doses of inactivated SARS-CoV-2 vaccines at 67 ± 6 years, respectively, indicating that the mRNA vaccines is more potent than the inactivated SARS-CoV-2 vaccines.^37^

The benefits of post-vaccination serologic testing outweigh the potential risks. Zitt et al. reported there were median titer of 1,440 BAU/mL in documented hepatitis B virus (HBV) vaccine responders (anti-HBs antibody ≥10 mIU/mL) and median titer of 308.5 BAU/mL in non-responders after two doses of mRNA vaccination (*P* = 0.035), suggesting that post-vaccination testing might predict the general immune competence.^36^ All anti-SARS-CoV-2 spike IgG-positive patients recovered from the infection respond well to the vaccine, which indirectly proves this phenomenon.^29-30^ If this theory turns out to be correct, then it is possible that SARS-CoV-2 vaccine responders have a strong ability to produce antibodies against variants through asymptomatic infections. This may support the Government-issued “immunity passports” to demonstrate an individual’s immune ability according to the WHO IS (≥ 50 BAU/mL) after recovered from COVID-19 or SARS-CoV-2 vaccination.

The most significant benefit of post-vaccination serologic testing is to save patient lives. Chukwu et al. reported clinical findings in a group of kidney transplant recipients vaccinated with 2 doses of vaccines (72% of BNT162b2, 28% of AZD1222). There were 22 breakthrough infections and 3 deaths after vaccination, including 77% (17/22) infections and 13.6% (3/22) deaths in the seronegative group and only 23% (5/22) infections and 0% (0/22) deaths in the seropositive group.^38^ However, this study did not use the WHO IS to get a cutoff for the responder. Therefore, there may be some non-responders (fold-index <4) in the seropositive group according to our study.

For SARS-CoV-2 vaccine non-responders, one benefit from post-vaccination serologic testing to the patient is to get a booster shot as soon as possible.^39-40^ For persistent non-responders to SARS-CoV-2 vaccination, anti-SARS-CoV-2 antibody injections could save these lives in the seronegative group after the vaccination.^41-43^

Another good example for post-vaccination serologic testing is the HBV vaccine. After the first hepatitis B vaccine was approved in the United States in 1981 and the recombinant hepatitis B vaccine developed by Maurice Hilleman was approved by the FDA in 1986, it took scientists more than 20 years to realize that the vaccine did not provide good protection for the elderly and certain immunocompromised populations and put them at risk of breakthrough infections after vaccination.^44-45^ Szmuness et al. have first reported that 7.4% of immunized individuals fail to elicit detectable specific antibodies after two doses of hepatitis B vaccine, suggesting that there are non-responders in the population in 1982.^46^ Roome et al. have found in 1993 that 11.9% of individuals with hepatitis B vaccine were no or inadequate levels of antibody, suggesting that post-vaccination testing should be done at intervals of 30 to 90 days after the last vaccine dose.^47^ Many subsequent studies have shown that the elderly and immunocompromised populations are associated with reduced vaccine responses to hepatitis B vaccination.^45-47^ The CDC has recommended post-vaccination serologic testing using the WHO IS for immunocompromised individuals following HBV vaccination.^45^ For persistent non-responders (anti-HBs antibody <10 mIU/mL, the WHO IS 07/164) to HBV vaccination, anti-HBV Ig injections are recommended if exposed to HBV.^45^

Furthermore, the lower immune cell count may form the major risk factor for non-responders after administration of full SARS-CoV-2 vaccine in our study. Van Oekelen et al. have demonstrated that 32.3% (10/31) of multiple myeloma patients with severe lymphopenia (<500/mm^3^) remained negative for SARS-CoV-2 spike IgG after two doses of mRNA vaccines (OR 2.89, 95% CI 1.10–7.20, *P* = 0.018).^48^ Similarly, two studies reported that 63.7–77.3% of patients who had a history of anti-CD20 therapy for B cell depletion remained negative for SARS-CoV-2 IgG after receiving mRNA vaccines, suggesting that B cells are required for humoral immunity following COVID-19 vaccines.^49-50^ Hence, further clinical trials must be performed to finalize effective booster shots for immunocompromised people after administering the complete dose in the general population.^51-55^

According to this study, the anti-spike IgG seropositivities were 99.7% and 98.9% in the 18–59 yr and ≥60 yr groups, respectively. Additionally, certain naïve immune cells, such as CD4 cells, CD8 cells, and B cells exhibited significant waning in the elderly people, suggesting that the non-seroconversion rates were higher in individuals with lower immune cell counts. Incidentally, the anti-spike IgG seroprevalence or positivity was inconsistent with seroconversion rates observed in our study, thereby suggesting that anti-spike IgG positivity might not be a suitable predictor for the seroconversion rates. Our data showed that 7.5-11.7% of non-responders existed in the population, even some non-responders with anti-spike IgG positivity, supporting the CDC’s concern that some non-responders are positive for anti-spike IgG after vaccination.^56^ An FDA EUA quantitative assay with the WHO IS (20/136) may help to address this issue.^26-27^

There are several potential strategies that can be employed to reduce the COVID-19 mortality rate below 0.13 % of that caused by influenza virus. These include the following measures: (1) Increase the vaccination rate of the population;^2^ (2) Develop vaccines against emerging and potential variants;^57-62^ (3) Administer booster vaccines for non-responders;^63-65^ (4) Assessment of humoral immune response of children, the elderly, and immunocompromised persons within 1–3 months after 4^th^ dose;^45, 65-70^ and (5) Incorporate additional protective measures for individuals with persistent (4^th^ or 5^th^ dose) negative humoral immune response after booster vaccination, such as injection of anti-SARS-CoV-2 immunoglobulins, antiviral drug treatment, usage of N95 masks in endemic areas, etc.^41-43, 71-74^

## CONCLUSIONS

Immune cells such as CD4 cells, CD8 cells, and B cells and anti-spike IgG levels were significantly reduced in the elderly. There were 7.5% non-responders among the 18–59 yr group and 11.7% in the ≥60 yr group. A titer of anti-SARS-CoV-2 spike IgG is blew 50 BAU/mL to be considered as non-responders at intervals of 30 to 90 days after the last vaccine dose. Booster vaccination may be recommended for non-responders to reduce the disease severity and mortality.

## Data Availability

All data produced in the present work are contained in the manuscript

## Limitations

Study limitations include small sample size, data was only sourced from a single center, and lack of RBD/spike-specific cellular immune assessments.

## Author Contributions

Drs Zeng and Xu had full access to all of the data in the study and take responsibility for the integrity of the data and the accuracy of the data analysis. Drs Zeng and Yang served as co–first authors and contributed equally to the work. Concept and design: All authors. Acquisition, analysis, or interpretation of data: All authors. Drafting of the manuscript: Zeng and Xu. Critical revision of the manuscript for important intellectual content: Zeng, Huang and Xu. Statistical analysis: Zeng, Huang and Xu. Obtained funding: Zeng and Gao. Administrative, technical, or material support: Zeng, Yang, Lin, Xu. Supervision: Zeng and Xu.

## Institutional Review Board Statement

The study was approved by the institutional ethics committee (No. S2021-481-01).

## Informed Consent Statement

All study participants provided their informed written consent.

## Data and materials availability

All data are available in the manuscript or the supplementary materials.

